# Advance Care Planning Documentation Completeness and End-of-Life Care: Trends and Associations Among U.S. Older Adults

**DOI:** 10.64898/2026.04.07.26350311

**Authors:** Zhigang Xie, Molly Jacobs, Jiaming Liang, Bhavana Patel, Young-Rock Hong

**Affiliations:** Department of Health Services Research, Management, and Policy, College of Public Health and Health Professions, University of Florida, Gainesville, Florida, USA; School of Public Health, University of Texas Health Science Center at Houston, San Antonio, Texas, USA; Department of Neurology, University of Florida College of Medicine, Gainesville, Florida, USA; Fixel Institute for Neurologic Diseases, University of Florida, Gainesville, Florida, USA; Department of Family and Preventive Medicine, Emory University School of Medicine, Atlanta, Georgia, USA; Winship Cancer Institute of Emory University, Atlanta, Georgia, USA

**Keywords:** Advance Care Planning, Living Will, Durable Power of Attorney, End-of-Life Care

## Abstract

**Background:** Advance care planning (ACP) documentation, including living wills and durable power of attorney (DPOA), is intended to support goal-concordant end-of-life care. However, it is unknown if comprehensive documentation confers additional benefits, and how these associations vary across clinical contexts.

**Methods:** We used 2010–2022 Health and Retirement Study exit interview data to examine associations between ACP documentation and end-of-life care among U.S. adults aged ≥50 years.

Documentation was categorized as none, one document (living will or DPOA), or two documents (both). Outcomes included intensive care unit (ICU) use, life-sustaining treatment, hospice enrollment, and out-of-hospital death. Modified Poisson regression models were used to estimate adjusted risk ratios (aRRs), and temporal trends in documentation were assessed using joinpoint regression.

**Results:** Among 5,622 decedents representing 23.2 million individuals, 42.7% had two documents and 28.9% had none, documentation increased substantially around 2014. Compared with no documentation, having any documentation was associated with lower likelihood of life-sustaining treatment (aRR=0.85, 95% CI: 0.74–0.98) and higher likelihood of hospice enrollment (aRR=1.43, 95% CI: 1.28–1.60) and out-of-hospital death (aRR=1.11, 95% CI: 1.06–1.18), but not ICU use. Having two documents showed similar patterns, with modest differences compared with one document after adjustment. Associations were stronger among decedents with expected death and attenuated among those with unexpected death.

**Conclusions:** Comprehensive ACP documentation is associated with less aggressive end-of-life care and greater hospice use, though the incremental benefits of two documents are modest. Findings highlight the importance of documentation within care planning processes and the clinical context.

## Introduction

Advance care planning (ACP) is central to align end-of-life care with patient preferences. ACP allows individuals to articulate their future medical treatment and designate surrogate decision-makers, thereby promoting care aligned with patients’ values and goals.^1–4^ Two key components of ACP include living wills, which usually document healthcare preferences such as life-sustaining treatments, and durable power of attorney (DPOA) for healthcare, which appoints a surrogate decision-maker to act on the patient’s behalf.^5^ Together, these comprehensive ACP documentations are intended to support shared decision-making and reduce uncertainty during critical clinical situations when patients may lack decisional capacity.^5–7^

Despite longstanding policy and clinical emphasis on ACP, evidence regarding its effectiveness in improving end-of-life outcomes remains mixed. Earlier studies suggested that ACP is associated with reduced use of aggressive interventions (e.g., intensive care unit [ICU] admission, mechanical ventilation, cardiopulmonary resuscitation) and increased hospice utilization.^8–13^ However, more recent research highlights substantial heterogeneity in these associations, with some studies finding limited or context-dependent effects, particularly after adjusting for patient preferences, illness trajectories, and healthcare system factors.^14–16^ For example, recent analyses using nationally representative datasets have shown that ACP documentation is associated with greater hospice use and a higher likelihood of dying in their preferred setting (e.g., outside of a hospital), but not with reductions in potentially burdensome care such as gastrostomy tube insertion, intubation/mechanical ventilation.^17^ One factor contributing to this heterogeneity is that most prior studies have operationalized ACP documentation as a binary construct (any vs. none), potentially obscuring important differences between partial and comprehensive ACP documentation.^18–20^ Emerging evidence emphasizes that ACP is an ongoing process rather than a one-time event, and that effective ACP requires integration of documentation with communication and clinical decision-making processes.^21–25^ In this context, having both a living will and a DPOA may represent a more complete and actionable form of ACP, potentially leading to more goal-concordant end-of-life care.^12^

Additionally, whether these additive associations differ by the anticipated trajectory of death is further unresolved question.^26^ When death is anticipated, there may be a greater opportunity for ACP to influence care decisions through communication and planning. In contrast, in cases of unexpected death (e.g., sudden death from a heart attack), the opportunity for ACP to shape care may be more limited, and the presence of a surrogate decision-maker may play a more critical role than documented preferences alone. However, to our knowledge, few prior studies explicitly examined whether the associations between ACP and end-of-life care differ by the expectedness of death.

To address these gaps, we used nationally representative data from the Health and Retirement Study (HRS) to examine additive associations between comprehensive ACP documentation (living wills and DPOA) and end-of-life care outcomes among U.S. older adults. We evaluated whether having both documents is associated with differences in intensive care use, life-sustaining treatment, hospice enrollment, and location of death, compared with having one or no documents. We further examined whether these associations vary by the expectedness of death and assessed temporal trends in ACP documentation. By distinguishing between partial and comprehensive ACP documentation, this study aims to provide new insights into the mechanisms by which ACP documentation may influence end-of-life care and to inform policies and interventions that promote goal-concordant end-of-life care.

## Methods

### Data Source and Study Population

We conducted a retrospective cross-sectional study using HRS data, a nationally representative longitudinal survey of U.S. adults aged 50 years and older.^27,28^ We analyzed exit interview data collected from 2010 to 2022, generally covering decedents who died one to three years before the interview. The interviews were administered to proxy respondents following a participant’s death and provide detailed information on end-of-life experiences, healthcare utilization, and advance care planning.^29,30^ The analytic sample included decedents aged 50 years or older with complete information on ACP documentation (living will and/or DPOA) and having positive sampling weights (n=6,986). After excluding respondents with missing data on sociodemographic and death-related variables, the main analytic sample consisted of 5,622 decedents, representing approximately 23.2 million individuals nationwide. The University of Florida Institutional Review Board determined that this study involved non-human subjects research using publicly available, de-identified data and was therefore exempt from review. This study followed the Strengthening the Reporting of Observational Studies in Epidemiology (STROBE) reporting guideline for cross-sectional studies.

### Measures

The primary exposure was the comprehensive ACP documentation completion status, based on proxy-reported information on whether the decedent had a living will and/or a DPOA for healthcare. We operationalized it using two approaches. First, a binary indicator distinguished individuals with any documentation (living will and/or DPOA) from those with none. Second, an ordinal variable categorized individuals into three mutually exclusive groups to ensure adequate cell sizes for multivariable regression analyses: no documentation, one document (either a living will or DPOA), and two documents (both a living will and DPOA). This categorization allowed examination of the additive associations of having both components of ACP documentation.

We examined four end-of-life care outcomes reported in the exit interview: ICU use in the last two years of life, use of life-sustaining treatments, hospice enrollment prior to death, and location of death categorized as out-of-hospital versus hospital death. All outcomes were treated as binary variables (yes, no).

Demographic characteristics included age at death (50–64, 65–84, ≥85 years), sex (male, female), race/ethnicity (non-Hispanic White, non-Hispanic Black, Hispanic, non-Hispanic Other), educational attainment (GED or lower, high school graduate, some college, college graduate or above), marital status (married or living with a partner, divorced or separated, widowed, never married), poverty status categorized into income quartiles (Q1–Q4), with Q1 representing the lowest income group, cause of death (cancer, cardiovascular disease, or other causes), and whether the death was expected at about the time it occurred (yes/no). These variables were selected based on prior literature demonstrating their associations with advance care planning and end-of-life decision-making.^31,32^

### Statistical Analysis

We first compared decedents’ characteristics by ACP documentation status using chi-square tests. We then estimated the annual prevalence of ACP documentation completion by year of death from 2008 to 2021 and assessed linear trends over time. These analyses were additionally stratified by expectedness of death to examine potential differences in temporal patterns. To further characterize trends, we conducted joinpoint regression analyses to estimate annual percent change (APC) in ACP documentation completion and to evaluate the statistical significance of changes in trend slopes across distinct time periods.^33^

Next, we calculated the prevalence of each end-of-life outcome across ACP documentation groups. To examine associations between ACP documentation and end-of-life care outcomes, we used modified Poisson regression models with robust variance estimation to estimate adjusted risk ratios (aRRs) and 95% confidence intervals (CIs) adjusting for aforementioned covariates.

This approach was selected to provide valid and interpretable estimates of relative risk for common binary outcomes.^34^ Models were specified using both binary (any vs. none) and ordinal (none, one document, two documents) exposure definitions, with no ACP documentation as the reference category. We further conducted stratified analyses by expected versus unexpected death to assess whether associations varied by the anticipated nature of death.

All analyses accounted for the HRS complex survey design, incorporating last wave interview sampling weights, strata, and primary sampling units to generate nationally representative estimates. Statistical analyses were performed using SAS version 9.4 (SAS Institute Inc.) and Stata version 19 (StataCorp), and statistical significance was defined as a two-sided p-value less than 0.05.

## Results

Overall, among the 5,622 decedents, 28.9% of decedents had no ACP documentation, 28.3% had one document, and 42.7% had two documents (**Table 1**). ACP documentation differed significantly across sociodemographic and clinical characteristics. Decedents with both documents were more likely to be older, with nearly half aged 85 years or older, compared with less than one-fifth among those without any documentation. They were also more likely to be female (53.5%), non-Hispanic White (90.2%), and have college or above educational attainment (23.8%). In addition, individuals with two documents were more likely to have experienced an expected death compared with those without documentation (66.1% vs. 43.6%).

**Table 1.**
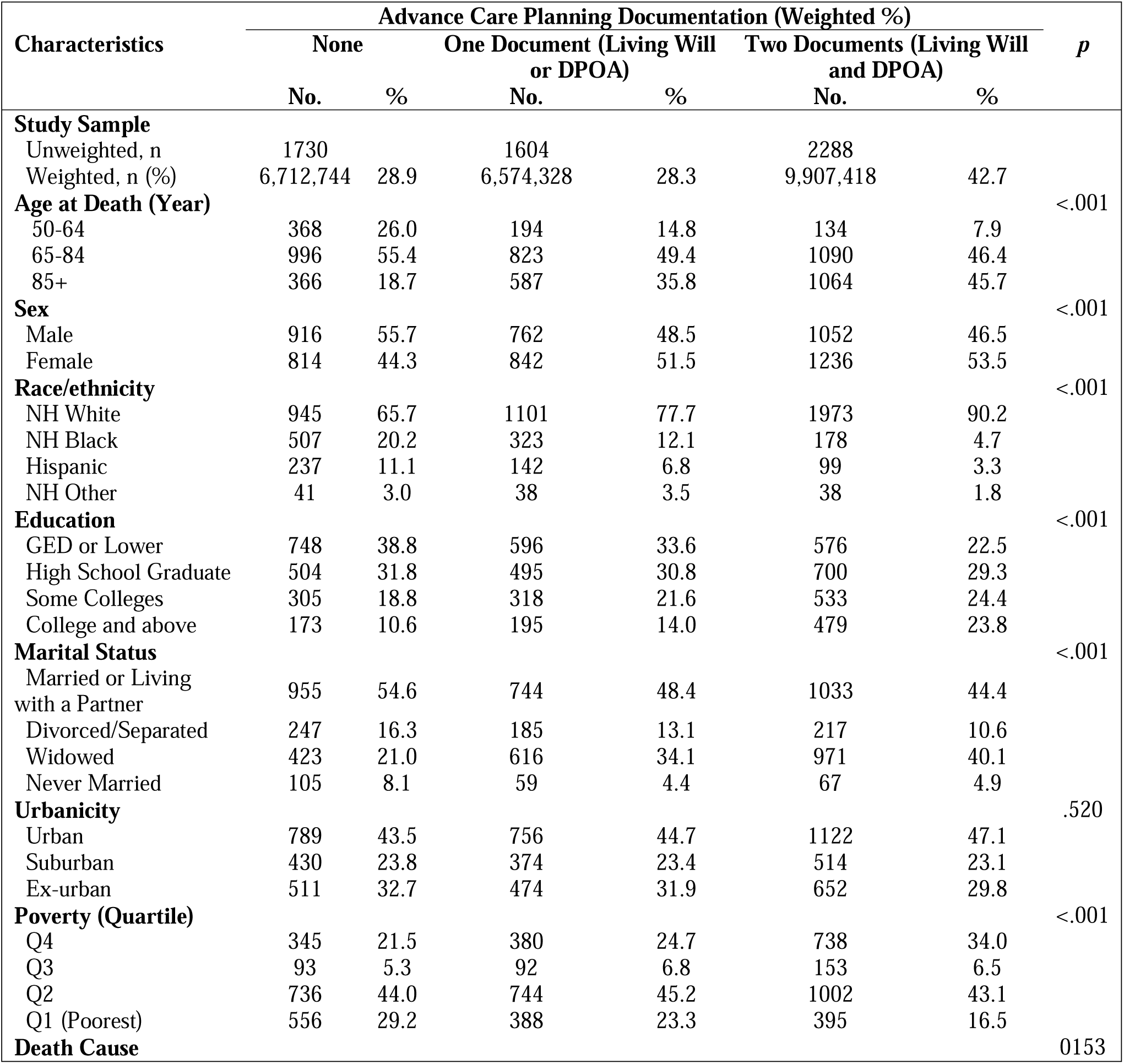
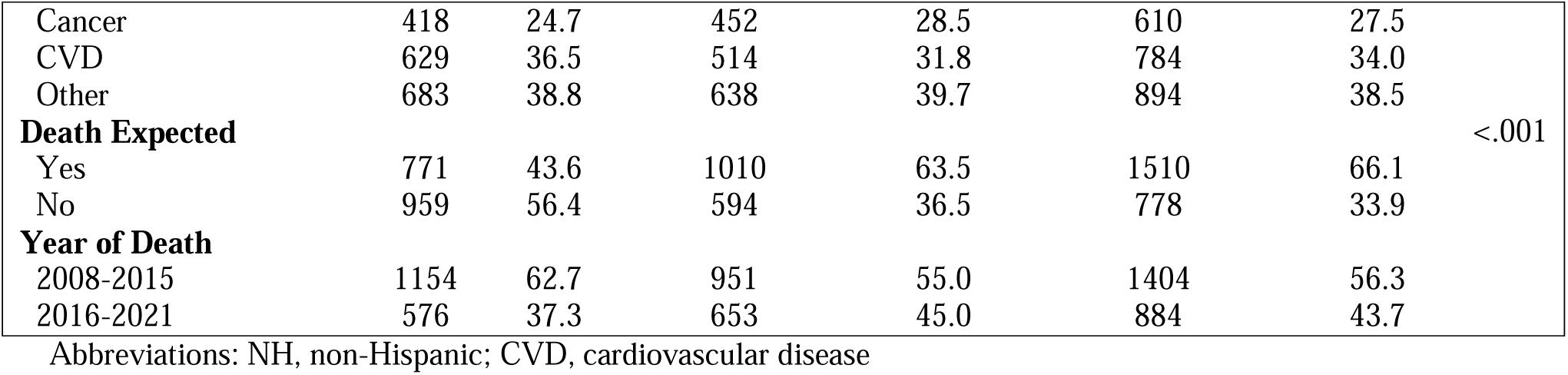
Respondent Reported Characteristics of Decedents by Dementia Status Before Death (n = 5,622, Weighted n = 23,194,490)

In unadjusted analyses, ACP documentation was consistently associated with less aggressive end-of-life care and greater use of hospice services (**Table 2**). ICU use was reported in 51.7% (95% CI: 48.6%-54.8%) of decedents without ACP documentation, compared with 48.3% (95% CI: 45.0%-51.7%) among those with one document and 45.0% (95% CI: 42.4%-47.7%) among those with both documents. A similar pattern was observed for life-sustaining treatment, with 38.9% (95% CI: 35.6%-42.3%) of those without ACP documentation receiving such care, compared with 28.1% (95% CI: 24.4%-31.8%) among those with one document and 24.3% (95% CI: 22.3%-26.3%) among those with both documents. In contrast, hospice enrollment was substantially higher among individuals with ACP documentation, particularly those with both documents, among whom 58.8% (95% CI: 55.4%-62.2%) received hospice care compared with 29.3% (95% CI: 26.2%-32.4%) among those without documentation. Similarly, out-of-hospital death was more common among those with ACP documentation, occurring in 73.2% (95% CI: 70.6%-75.8%) of decedents with both documents compared with 59.1%(95% CI: 56.1%-62.1%) among those with none.

**Table 2.** Prevalence and Relative Risks of End-of-Life Care Outcomes Among Decedents Using SurveylJWeighted Modified Poisson Regression.

| Characteristics | Prevalence |  |  | Regression |  |  |
| --- | --- | --- | --- | --- | --- | --- |
|  | % | 95% CI |  | p | aRR* | 95% CI |
| Intensive Care Unit Use (n =4,482, Weighted n =18,354,145) |  |  |  |  |  |  |
| ACP Documentation (Binary) |  |  |  | .004 |  |  |
| None | 51.7 | 48.6 | 54.8 |  | 1.00 |  |
| Any (Living Will and/or DPOA) | 46.4 | 44.4 | 48.3 |  | 1.03 | 0.96 1.10 |
| ACP Documentation (Ordinal) |  |  |  | .008 |  |  |
| None | 51.7 | 48.6 | 54.8 |  | 1.00 |  |
| One Document (Living Will or DPOA) | 48.3 | 45.0 | 51.7 |  | 1.04 | 0.96 1.13 |
| Two Document (Living Will and DPOA) | 45.0 | 42.4 | 47.7 |  | 1.02 | 0.93 1.11 |
| Life-Support Use (n = 4,470, Weighted n=18,328,959) |  |  |  |  |  |  |
| ACP Documentation (Binary) |  |  |  | <.001 |  |  |
| None | 38.9 | 35.6 | 42.3 |  | 1.00 |  |
| Any (Living Will and/or DPOA) | 25.8 | 23.8 | 27.8 |  | 0.85 | 0.74 0.98 |
| ACP Documentation (Ordinal) |  |  |  | <.001 |  |  |
| None | 38.9 | 35.6 | 42.3 |  | 1.00 |  |
| One Document (Living Will or DPOA) | 28.1 | 24.4 | 31.8 |  | 0.87 | 0.75 1.02 |
| Two Document (Living Will and DPOA) | 24.3 | 22.3 | 26.3 |  | 0.84 | 0.71 0.98 |
| Hospice Use Before Death (n = 3,679, Weighted n=16,005,182) |  |  |  |  |  |  |
| ACP Documentation (Binary) |  |  |  | <.001 |  |  |
| None | 29.3 | 26.2 | 32.4 |  | 1.00 |  |
| Any (Living Will and/or DPOA) | 55.9 | 53.2 | 58.7 |  | 1.43 | 1.28 1.60 |
| ACP Documentation (Ordinal) |  |  |  | <.001 |  |  |
| None | 29.3 | 26.2 | 32.4 |  | 1.00 |  |
| One Document (Living Will or DPOA) | 51.6 | 47.7 | 55.6 |  | 1.37 | 1.20 1.55 |
| Two Document (Living Will and DPOA) | 58.8 | 55.4 | 62.2 |  | 1.48 | 1.32 1.66 |
| Out-Of-Hospital Death (n = 5,619, Weighted n=23,182,189) |  |  |  |  |  |  |
| ACP Documentation (Binary) |  |  |  | <.001 |  |  |
| None | 59.1 | 56.1 | 62.1 |  | 1.00 |  |
| Any (Living Will and/or DPOA) | 72.3 | 70.4 | 74.3 |  | 1.11 | 1.06 1.18 |
| ACP Documentation (Ordinal) |  |  |  | <.001 |  |  |
| None | 59.1 | 56.1 | 62.1 |  | 1.00 |  |
| One Document (Living Will or DPOA) | 71.0 | 68.5 | 73.6 |  | 1.11 | 1.05 1.17 |
| Two Document (Living Will and DPOA) | 73.2 | 70.6 | 75.8 |  | 1.12 | 1.05 1.19 |

Abbreviation: ACP, advance care planning; aRR, adjusted risk ratio
\*Models were adjusted for age at death, sex, race/ethnicity, educational attainment, marital status, urbanicity, poverty status, cause of death, expectedness of death, and year of death.

After adjustment for sociodemographic and clinical covariates, having any ACP documentation was associated with significantly lower likelihood of life-sustaining treatment (aRR=0.85, 95% CI: 0.74–0.98) and higher likelihood of hospice enrollment (aRR=1.43, 95% CI: 1.28–1.60) and out-of-hospital death (aRR=1.11, 95% CI: 1.06–1.18) (**Table 2**). There was no statistically significant association between having any ACP documentation and ICU use.

When the exposure was modeled ordinally, an additive pattern was observed, although differences between one and two documents were attenuated in adjusted models. Compared with having no ACP documentation, having two documents was associated with a lower likelihood of life-sustaining treatment (aRR=0.84, 95% CI: 0.71–0.98), as well as higher likelihood of hospice enrollment (aRR=1.48, 95% CI: 1.32–1.66) and out-of-hospital death (aRR=1.12, 95% CI: 1.05–1.19). Although point estimates generally suggested less aggressive care patterns among those with both documents compared with one document, these differences were modest and not consistently statistically significant after adjustment.

In stratified analyses (**Table 3**), associations between ACP documentation and end-of-life care varied by expectedness of death. Among decedents whose death was expected, ACP documentation were associated with lower likelihood of life-sustaining treatment and higher likelihood of hospice enrollment and out-of-hospital death. Specifically, having both documents was associated with a 19% (aRR=0.81, 95% CI: 0.66–0.99) lower likelihood of life-sustaining treatment, a 35% (aRR=1.35, 95% CI: 1.21–1.51) higher likelihood of hospice use, and a 20% (aRR=1.20, 95% CI: 1.10–1.30) higher likelihood of out-of-hospital death compared with having no ACP documentation. In contrast, among decedents with unexpected death, associations were generally attenuated, particularly for ICU use and out-of-hospital death. However, ACP documentation remained strongly associated with hospice enrollment even in this group, with those having both documents demonstrating substantially higher likelihood of hospice use compared with those without documentation (aRR=1.88, 95% CI: 1.29–2.74). Across both strata, there were no significant associations between ACP documentation and ICU use after adjustment.

**Table 3.** Relative Risks of End-of-Life Care Outcomes Estimated via Modified Poisson Regression, Stratified by Death Expectedness.

| Characteristics | Expected Death |  |  | Unexpected Death |  |  |
| --- | --- | --- | --- | --- | --- | --- |
|  | aRR* | 95% CI |  | aRR* | 95% CI |  |
| Intensive Care Unit Use |  |  |  |  |  |  |
| ACP Documentation (Binary) |  |  |  |  |  |  |
| None | 1.00 |  |  | 1.00 |  |  |
| Any (Living Will and/or DPOA) | 0.96 | 0.85 | 1.08 | 1.08 | 0.98 | 1.18 |
| ACP Documentation (Ordinal) |  |  |  |  |  |  |
| None | 1.00 |  |  | 1.00 |  |  |
| One Document (Living Will or DPOA) | 0.97 | 0.84 | 1.11 | 1.10 | 0.98 | 1.24 |
| Two Document (Living Will and DPOA) | 0.95 | 0.83 | 1.09 | 1.05 | 0.93 | 1.19 |
| Life-Support Use |  |  |  |  |  |  |
| ACP Documentation (Binary) |  |  |  |  |  |  |
| None | 1.00 |  |  | 1.00 |  |  |
| Any (Living Will and/or DPOA) | 0.81 | 0.68 | 0.96 | 0.86 | 0.74 | 1.00 |
| ACP Documentation (Ordinal) |  |  |  |  |  |  |
| None | 1.00 |  |  | 1.00 |  |  |
| One Document (Living Will or DPOA) | 0.81 | 0.67 | 0.99 | 0.90 | 0.75 | 1.07 |
| Two Document (Living Will and DPOA) | 0.81 | 0.66 | 0.99 | 0.82 | 0.67 | 1.00 |
| Hospice Use Before Death |  |  |  |  |  |  |
| ACP Documentation (Binary) |  |  |  |  |  |  |
| None | 1.00 |  |  | 1.00 |  |  |
| Any (Living Will and/or DPOA) | 1.30 | 1.16 | 1.46 | 1.80 | 1.29 | 2.49 |
| ACP Documentation (Ordinal) |  |  |  |  |  |  |
| None | 1.00 |  |  | 1.00 |  |  |
| One Document (Living Will or DPOA) | 1.25 | 1.09 | 1.42 | 1.72 | 1.22 | 2.41 |
| Two Document (Living Will and DPOA) | 1.35 | 1.21 | 1.51 | 1.88 | 1.29 | 2.74 |
| Out-Of-Hospital Death |  |  |  |  |  |  |
| ACP Documentation (Binary) |  |  |  |  |  |  |
| None | 1.00 |  |  | 1.00 |  |  |
| Any (Living Will and/or DPOA) | 1.19 | 1.10 | 1.29 | 1.08 | 0.98 | 1.19 |
| ACP Documentation (Ordinal) |  |  |  |  |  |  |
| None | 1.00 |  |  | 1.00 |  |  |
| One Document (Living Will or DPOA) | 1.17 | 1.08 | 1.28 | 1.09 | 0.99 | 1.21 |
| Two Document (Living Will and DPOA) | 1.20 | 1.10 | 1.30 | 1.07 | 0.96 | 1.20 |

Abbreviation: ACP, advance care planning; aRR, adjusted risk ratio
\*Models were adjusted for age at death, sex, race/ethnicity, educational attainment, marital status, urbanicity, poverty status, cause of death, and year of death.

Temporal trends indicated a substantial increase in any ACP documentation, either Living will or DPOA, and two documents over time (p<.05), with overall higher prevalence in the expected death group than unexpected death group (**Figure 1**). The prevalence of any ACP documentation remained relatively stable between 2008 and 2016 but increased significantly thereafter, reaching more than 86% by 2021. Joinpoint analysis identified a significant inflection point around 2014, after which the prevalence increased at an annual rate of approximately 2.7% (**Figure 2**). Similar patterns were observed for durable power of attorney and completion of both documents, with evidence of a reversal from declining or stable trends prior to approximately 2012 to significant increases thereafter. These findings suggest a growing uptake of advance care planning, particularly comprehensive documentation involving both treatment preferences and surrogate designation.

**Figure 1.**
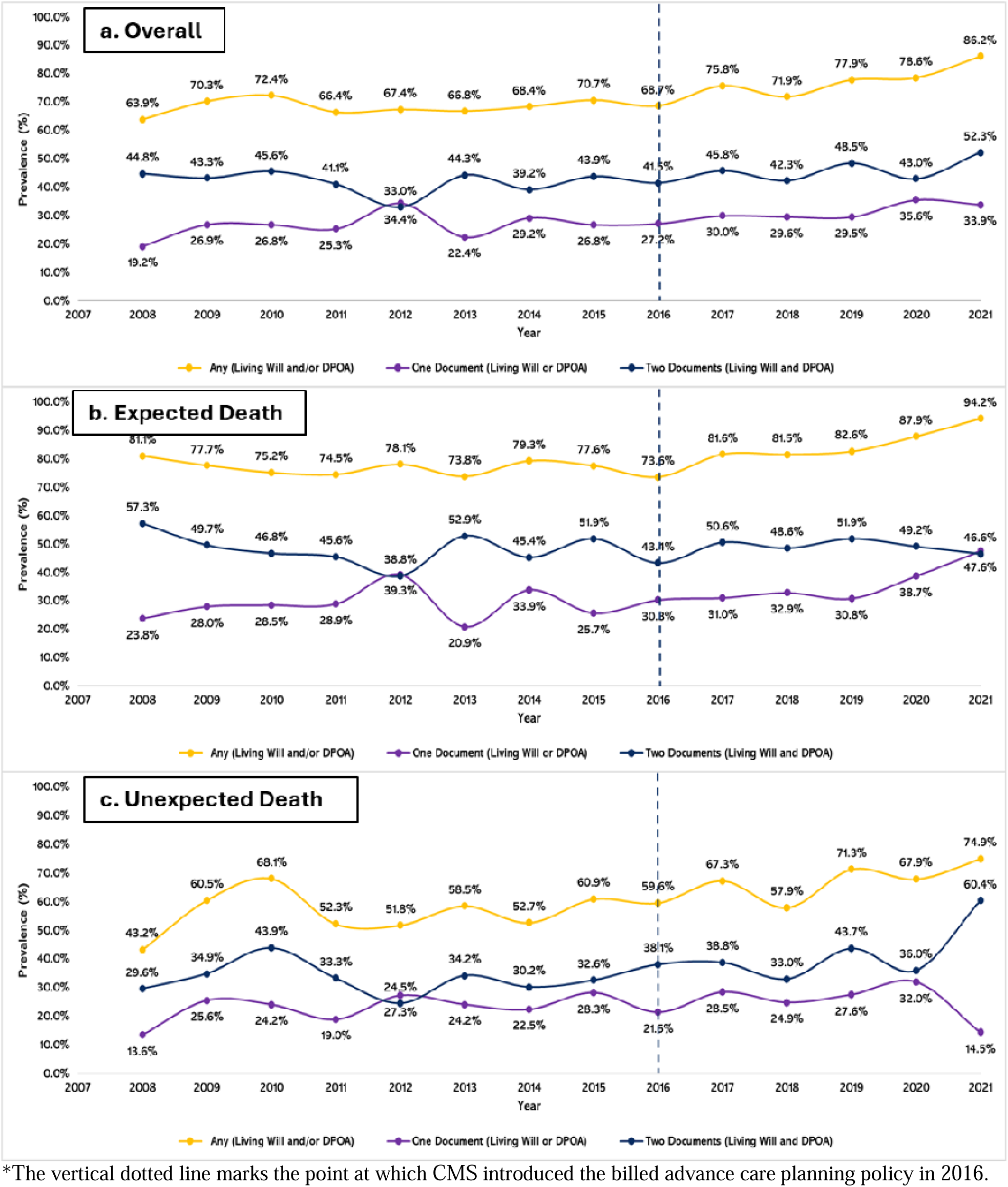
**Trends in advance care planning documentation (living wills and durable power of attorney [DPOA]) before death among decedents, 2008–2021**

**Figure 2.**
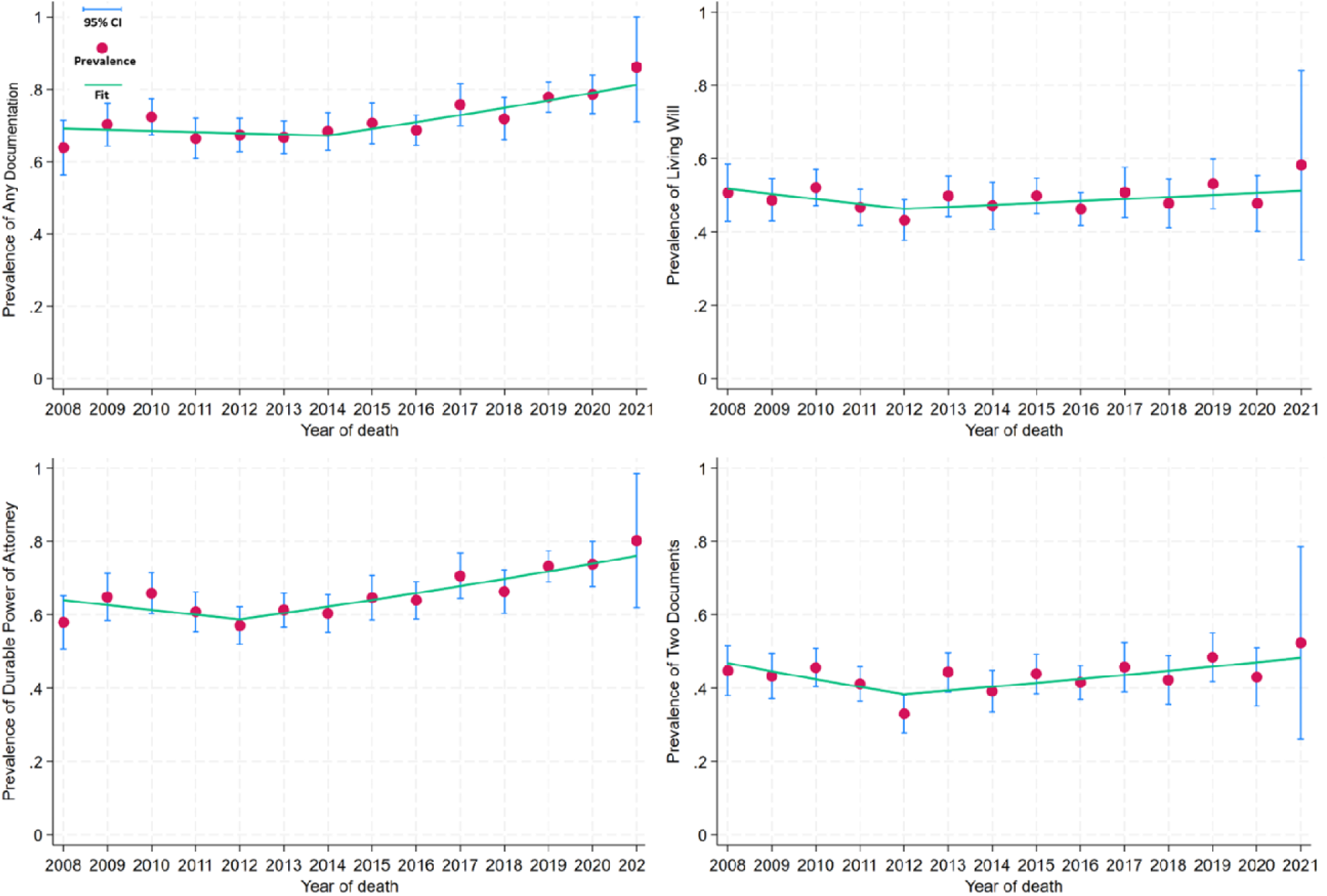
Jointpoint analysis of any advance care planning documentation (living wills and/or durable power of attorney [DPOA]), living will, DPOA, and two documents (living will and DPOA) before death among decedents, 2008–2021. The figure displays the survey-weighted prevalence of any advance care planning documentation (living will / durable power of attorney [DPOA]), living will, DPOA, and two documents (living will and DPOA) among decedents by year of death from 2008 to 2021. Blue dots represent the observed annual weighted prevalence, while the red line represents the fitted segmented regression trend from the Joinpoint model. **Any advance care planning documentation:** joinpoint analysis identified a trend change around 2014. Between 2008 and 2014, the prevalence remained relatively stable, fluctuating between 63.9% and 72.4%. After 2014, the prevalence increased steadily, with an estimated annual percentage change (APC) of approximately 2.7%, reaching over 86.2% by 2021 (p=0.003). **Living will:** joinpoint analysis suggested a change in trend around 2012. The prevalence declined modestly between 2008 and 2012 (APC, −3.1% per year) and increased thereafter (APC, 3.1% per year), although neither trend reached statistical significance. **DPOA**: joinpoint analysis identified a change in trend around 2012. The prevalence declined modestly between 2008 and 2012 (APC, −2.1%; p=0.11) and increased significantly thereafter (APC, 2.9%; p<0.001). The change in slope at the joinpoint was statistically significant (p=0.007), indicating a shift from a declining to an increasing trend. **Two documents**: joinpoint analysis identified a significant change in trend around 2012. The prevalence declined between 2008 and 2012 (APC, −4.9%; p=0.042) but increased thereafter (APC, 2.6%; p=0.029). The change in slope at the joinpoint was statistically significant (p=0.023), indicating a reversal in the trend beginning in 2012.

## Discussion

In this nationally representative study of U.S. older adult decedents, we observed a substantial increase in ACP documentation over time, particularly after a joinpoint inflection around 2014. Having any ACP documentation was associated with more favorable end-of-life outcomes compared with having no documentation, though the incremental benefit of having both a living will and a DPOA relative to a single document was modest after covariate adjustment. In stratified analyses, associations with life-sustaining treatment and out-of-hospital death were more pronounced among individuals with expected death and attenuated among those with unexpected death; however, ACP documentation remained strongly associated with hospice enrollment regardless of death expectedness. Together, these findings suggest that while ACP documentation is associated with less aggressive end-of-life care patterns, much of the observed benefit may be driven by broader processes underlying ACP rather than the accumulation of documents alone

### Temporal trend in ACP documentation

The observed increase in ACP documentation prevalence over time, particularly the acceleration identified by joinpoint analysis around 2014, likely reflects a confluence of policy, clinical, and cultural shifts that have elevated the prominence of advance care planning in routine care. The introduction of Medicare reimbursement for ACP discussions in 2016, alongside growing emphasis on patient-centered care and serious illness communication, may have incentivized clinicians to engage patients in these conversations and formalize documentation.^21,23^ In addition, broader public awareness of end-of-life planning that was driven by initiatives such as The Conversation Project and increased attention to goal-concordant care may have contributed to rising uptake. Despite these encouraging trends, prior work suggests that disparities in ACP completion persist across racial, socioeconomic, and educational groups,^20^ which is consistent with the sociodemographic gradients observed in our sample. Taken together, these findings suggest that while ACP is becoming more common, equitable access to and engagement in ACP remain important areas for intervention.

### Modest incremental benefit of having both documents

Although having both a living will and a durable power of attorney represents a more comprehensive form of ACP, our findings indicate that the incremental benefit of dual documentation (beyond having a single document) is modest after adjustment for sociodemographic and clinical factors. This pattern aligns with emerging conceptualizations of ACP as a dynamic, communication-driven process rather than a static documentation event.^25,35,36^ The presence of any documentation may serve as a proxy for underlying patient engagement, preferences for less aggressive care, or prior discussions with clinicians and family members, which may be the primary drivers of observed differences in end-of-life outcomes. In this context, the marginal gains associated with additional documentation may reflect diminishing returns when preferences are already known or when surrogate decision-makers are sufficiently prepared. These findings are consistent with prior studies demonstrating that ACP’s effectiveness depends not only on the existence of documents but also on their integration into clinical workflows and real-time decision-making. ^14,17,37^Accordingly, interventions focused solely on increasing documentation rates may have a limited impact unless paired with efforts to enhance communication quality, surrogate preparedness, and accessibility of ACP information at the point of care.

### Variation by expectedness of death

The stronger associations between ACP documentation and life-sustaining treatment, hospice enrollment, and out-of-hospital death observed among decedents with expected death are consistent with the premise that anticipated dying trajectories create greater opportunity for iterative discussions, prognostic awareness, and alignment of care with patient preferences.^38,39^ In cases of unexpected death, such as acute events or rapid clinical deterioration, the window for ACP to influence care may be limited, and decisions may be driven more by emergent clinical circumstances than by previously documented preferences. A notable exception to the general attenuation pattern was hospice enrollment, for which ACP documentation showed its strongest association in those with unexpected death. One interpretation is that among decedents whose deaths were expected, hospice enrollment is relatively common regardless of documentation status, leaving less room for documentation to differentiate care patterns. In contrast, when death is unexpected, documented preferences or a designated surrogate may function as a more decisive signal that redirects care toward hospice even in acute clinical situations where extended deliberation is not feasible. Our findings suggest that the mechanisms through which ACP documentation influences hospice referral differently from other end-of-life care decisions; surrogate activation and the documentation signaling may be more important than preference specification alone when death is not expected.^40^ Further investigation of these pathway-specific effects is warranted.

### Policy Implications

These findings have important implications for policy and clinical practice. While recent policy efforts such as Medicare reimbursement for advance care planning have likely contributed to increased documentation,^21,38,41,42^ our results suggest that expanding documentation alone may be insufficient to meaningfully improve end-of-life care. Instead, policies should prioritize the quality and integration of ACP processes, including structured, iterative communication between patients, families, and clinicians, as well as preparation and support for surrogate decision-makers. Incorporating standardized measures of ACP communication quality into value-based care frameworks may help ensure that documentation reflects informed, preference-sensitive decision-making rather than a procedural requirement. In addition, improving interoperability and accessibility of ACP documentation within electronic health records across care settings is critical to ensuring that patient preferences are available and actionable at the point of care, particularly during acute transitions. Together, these strategies may enhance ACP’s capacity to promote goal-concordant care beyond mere documentation.

## Limitations

These findings should be interpreted in light of the following limitations. First, the use of proxy-reported exit interview data may introduce recall bias or misclassification, particularly for ACP documentation and end-of-life care experiences. Although proxy reports are widely used in end-of-life research and are often the only feasible source of information after death, their accuracy may vary depending on the respondent’s knowledge and involvement in care. Second, our measures of ACP documentation capture the presence of living wills and durable power of attorney but do not reflect the content, quality, timing, or accessibility of these documents, nor whether they were available or adhered to at the point of care. As a result, we were unable to assess whether care was truly goal concordant. Third, outcomes were measured as broad indicators of healthcare utilization and may not fully capture the complexity of end-of-life decision-making or the appropriateness of care in specific clinical contexts. Fourth, stratification by expectedness of death was based on proxy assessment and may be subject to misclassification, particularly in cases with uncertain illness trajectories. Finally, the observational study design precludes causal inference. Individuals with ACP documentation may differ systematically from those without in ways not fully captured by measured covariates, such as health literacy, preferences for care, or engagement with the healthcare system, potentially leading to residual confounding. Despite these limitations, this study leverages a large, nationally representative dataset and robust analytic methods to provide new insights into the role of ACP documentation components in shaping end-of-life care.

## Conclusions

Comprehensive ACP documentation is associated with less aggressive end-of-life care and greater use of hospice services among U.S. older adults, with modest additional benefit observed for having both a living will and a DPOA compared with having a single document. Although we cannot directly measure goal-concordant care, the observed patterns (reduced life-sustaining treatments and increased hospice use) are consistent with care better aligned with patient preferences. The increasing prevalence of ACP documentation over time is encouraging; however, the general attenuation of associations in the context of unexpected death underscores the need to strengthen real-time decision-making support, including surrogate preparedness and clinician communication. Future work should examine whether earlier ACP (e.g., prior to functional decline) yields stronger effects than late-stage documentation.

## Acknowledgements

None.

## Author Contributions

**ZX:** Conceptualization; methodology; software; formal analysis; original draft; writing – review and editing.

**MJ:** Conceptualization; original draft; supervising; writing – review and editing; corresponding author.

**JL:** Conceptualization; writing – review and editing.

**BP:** Conceptualization; writing – review and editing.

**YH:** Conceptualization; methodology; supervising; writing – review and editing; senior author. **Ethical Considerations:** As the HRS is a publicly available, de-identified dataset, the Institutional Review Board (IRB) at the first author’s university determined that this study was non-human subject research.

## Consent to Participate

It is not applicable as this is a secondary data analysis with publicly available deidentified data.

## Consent for Publication

Not applicable.

## Data Availability Statement

The data supporting the findings of this study are openly available on the Health and Retirement Study website. These data were derived from the following resources available in the public domain: https://hrs.isr.umich.edu/data-products.

## References

1. McMahan RD, Tellez I, Sudore RL. Deconstructing the complexities of advance care planning outcomes: what do we know and where do we go? A scoping review. J Am Geriatr Soc. 2021;69(1):234–244.

2. Murray L, Butow PN. Advance care planning in motor neuron disease: A systematic review. Palliative & supportive care. 2016;14(4):411–432.

3. Fried TR, Bullock K, Iannone L, O’leary JR. Understanding advance care planning as a process of health behavior change. J Am Geriatr Soc. 2009;57(9):1547–1555.

4. Sudore RL, Lum HD, You JJ, et al. Defining advance care planning for adults: a consensus definition from a multidisciplinary Delphi panel. J Pain Symptom Manage. 2017;53(5):821–832. e1.

5. Sedini C, Biotto M, Crespi Bel’skij LM, Moroni Grandini RE, Cesari M. Advance care planning and advance directives: an overview of the main critical issues. Aging clinical and experimental research. 2022;34(2):325–330.

6. Rosca A, Karzig-Roduner I, Kasper J, Rogger N, Drewniak D, Krones T. Shared decision making and advance care planning: a systematic literature review and novel decision-making model. BMC Medical Ethics. 2023;24(1):64.

7. Hamilton IJ. Advance care planning in general practice: promoting patient autonomy and shared decision making. The British Journal of General Practice. 2017;67(656):104.

8. Ashana DC, Chen X, Agiro A, et al. Advance care planning claims and health care utilization among seriously ill patients near the end of life. JAMA network open. 2019;2(11):e1914471.

9. Crooks J, Rizk N, Simpson-Greene C, et al. Evaluating outcomes of advance care planning interventions for adults living with advanced illness and people close to them: A systematic meta-review. Palliat Med. 2025:02692163251344428.

10. Song K, Amatya B, Voutier C, Khan F. Advance care planning in patients with primary malignant brain tumors: a systematic review. Frontiers in oncology. 2016;6:223.

11. Walczak A, Butow PN, Bu S, Clayton JM. A systematic review of evidence for end-of-life communication interventions: who do they target, how are they structured and do they work? Patient Educ Couns. 2016;99(1):3–16.

12. Silveira MJ, Kim SY, Langa KM. Advance directives and outcomes of surrogate decision making before death. N Engl J Med. 2010;362(13):1211–1218.

13. Wright AA, Zhang B, Ray A, et al. Associations between end-of-life discussions, patient mental health, medical care near death, and caregiver bereavement adjustment. JAMA. 2008;300(14):1665–1673.

14. Jimenez G, Tan WS, Virk AK, Low CK, Car J, Ho AHY. Overview of systematic reviews of advance care planning: summary of evidence and global lessons. J Pain Symptom Manage. 2018;56(3):436–459. e25.

15. Pimsen A, Kao C, Hsu S, Shu B. The effect of advance care planning intervention on hospitalization among nursing home residents: a systematic review and meta-analysis. Journal of the American Medical Directors Association. 2022;23(9):1448–1460. e1.

16. Wang J, Zhou A, Peng H, et al. Effects of advance care planning on end-of-life decisions among community-dwelling elderly people and their relatives: a systematic review and meta-analysis. Annals of Palliative Medicine. 2023;12(3):57183–57583.

17. Wolff JL, Scerpella D, Giovannetti ER, et al. Advance care planning, end-of-life preferences, and burdensome care: a pragmatic cluster randomized clinical trial. JAMA internal medicine. 2025;185(2):162–170.

18. Zhu Y, Enguidanos S. Advance directives completion and hospital out of pocket expenditures. Journal of hospital medicine. 2022;17(6):437–444.

19. Wang Z, Huang Y, Zhang T, Wang Y, Sun S, Tao A. Association between Advance Directives and Invasive Life Support Use at End of Life: A Retrospective Cohort Study. J Pain Symptom Manage. 2026.

20. Yadav KN, Gabler NB, Cooney E, et al. Approximately one in three US adults completes any type of advance directive for end-of-life care. Health Aff. 2017;36(7):1244–1251.

21. Luth EA, Manful A, Weissman JS, et al. Practice billing for Medicare advance care planning across the USA. Journal of general internal medicine. 2022;37(15):3869–3876.

22. Gupta A, Jin G, Reich A, et al. Association of billed advance care planning with end of life care intensity for 2017 Medicare decedents. J Am Geriatr Soc. 2020;68(9):1947–1953.

23. Mehta A, Kelley AS. Advance care planning codes—getting paid for quality care. JAMA internal medicine. 2019;179(6):830–831.

24. Dingfield LE, Kayser JB. Integrating advance care planning into practice. Chest. 2017;151(6):1387–1393.

25. Hickman SE, Lum HD, Walling AM, Savoy A, Sudore RL. The care planning umbrella: the evolution of advance care planning. J Am Geriatr Soc. 2023;71(7):2350.

26. Wang X, Huang X, Wang W, Liao L. Advance care planning for frail elderly: are we missing a golden opportunity? A mixed-method systematic review and meta-analysis. BMJ open. 2023;13(5):e068130.

27. Juster FT, Suzman R. An overview of the Health and Retirement Study. J Hum Resour. 1995:S7–S56.

28. Fisher GG, Ryan LH. Overview of the health and retirement study and introduction to the special issue. Work, aging and retirement. 2018;4(1):1–9.

29. Health and Retirement Study, (RAND HRS Longitudinal File 2022 (V1)) public use dataset. Produced and distributed by the University of Michigan with funding from the National Institute on Aging (grant numbers NIA U01AG009740 and NIA R01AG073289). Accessed March 11, 2026.

30. RAND HRS Longitudinal File 2022 (V1) Produced by the RAND Center for the Study of Aging, with funding from the National Institute on Aging and the Social Security Administration. Accessed March 11, 2026.

31. Palmer MK, Jacobson M, Enguidanos S. Advance Care Planning For Medicare Beneficiaries Increased Substantially, But Prevalence Remained Low: Study examines Medicare outpatient advance care planning claims and prevalence. Health Aff. 2021;40(4):613–621.

32. Virdun C, Luckett T, Davidson PM, Phillips J. Dying in the hospital setting: a systematic review of quantitative studies identifying the elements of end-of-life care that patients and their families rank as being most important. Palliat Med. 2015;29(9):774–796.

33. Xu J, Lin Y, Yang M, Zhang L. Statistics and pitfalls of trend analysis in cancer research: a review focused on statistical packages. Journal of Cancer. 2020;11(10):2957.

34. Zou G. A modified poisson regression approach to prospective studies with binary data. Am J Epidemiol. 2004;159(7):702–706.

35. Sudore R, Boscardin J, Barnes D. A patient-facing advance care planning (ACP) website called PREPARE increases ACP documentation and engagement in a randomized trial of diverse older primary care patients at a VA medical center (TH307B). J Pain Symptom Manage. 2017;53(2):317–318.

36. Hickman SE, Lum HD, Unroe KT. Family caregiver perspectives on advance care planning discussions for residents with dementia led by trained nursing home staff: Insights from the APPROACHES project. Journal of the American Medical Directors Association. 2026:106154.

37. Jimenez G, Tan WS, Virk AK, Low CK, Car J, Ho AHY. State of advance care planning research: a descriptive overview of systematic reviews. Palliative & supportive care. 2019;17(2):234–244.

38. Ladin K, Bronzi OC, Gazarian PK, et al. Understanding The Use Of Medicare Procedure Codes For Advance Care Planning: A National Qualitative Study: Study examines the use of Medicare procedure codes for advance care planning. Health Aff. 2022;41(1):112–119.

39. Vellani S, Green E, Kulasegaram P, Sussman T, Wickson-Griffiths A, Kaasalainen S. Interdisciplinary staff perceptions of advance care planning in long-term care homes: a qualitative study. BMC palliative care. 2022;21(1):127.

40. Prater LC, Wickizer T, Bower JK, Bose-Brill S. The impact of advance care planning on end-of-life care: do the type and timing make a difference for patients with advanced cancer referred to hospice? American Journal of Hospice and Palliative Medicine®. 2019;36(12):1089–1095.

41. Weissman JS, Gazarian P, Reich A, et al. Recent trends in the use of Medicare advance care planning codes. J Palliat Med. 2020;23(12):1568–1570.

42. Belanger E, Loomer L, Teno JM, Mitchell SL, Adhikari D, Gozalo PL. Early utilization patterns of the new Medicare procedure codes for advance care planning. JAMA internal medicine. 2019;179(6):829–830.

